# Clinical genetics and its adjacent regimes

**DOI:** 10.1101/2020.06.04.20102939

**Authors:** T. Lange, T. Rigter, T. Vrijenhoek

**Affiliations:** Department of Genetics, University Medical Centre Utrecht, Utrecht, The Netherlands; Department of Clinical Genetics, Section Community Genetic & Amsterdam Public Health research institute, Amsterdam University Medical Center, Location VUmc, Amsterdam, The Netherlands

## Abstract

Clinical genetics is the prime application of genetics in healthcare, providing highly advanced and reliable diagnostics for patients with (mostly rare) disease of genetic origin. Whereas many novel technologies have expanded the genetic toolkit, integration or alignment with other areas of healthcare is often challenging. We hypothesise that this is due to the characteristics inherent to the regimes in which the genetic technologies were to be implemented. In order to facilitate integration of genetic applications in a rebooting and perhaps transforming healthcare system, we here provide insights in discrepancies between clinical genetics and four of its *adjacent regimes*; public health, human genetic research, non-genetic healthcare, and society. We conducted twelve semi-structured group interviews and a focus group to collect information on overlapping and distinctive elements of each regime. We identified three aspects in which the adjacent regimes differed considerably compared to clinical genetics; perception of data, expectations from technologies, and compartimentalisation units. Strikingly, divergence within each of these aspects was determined by elements of culture, and not – as is often thought – by elements of structure, e.g. regulation and policy. We conclude that implementation of genetics requires transdisciplinary empathy – understanding of the way of organizing, thinking and doing in adjacent regimes.

## Introduction

For two decades now, genetics has been destined to transform mainstream medicine^1–6^. To a certain extend it has, considering the elaborate genetic toolkit with highly advanced technologies and methodologies such as next-generation sequencing (NGS), genome-wide association studies (GWAS) and Clustered Regularly Interspaced Short Palindromic Repeats-associated nuclease 9 (CRIPR/Cas9). Still, the expected transformation of genetics into a widely used solution to support prevention, treatment and cure of many diseases has not occurred. Now – in a healthcare system that is gradually rebooting – there is an opportunity to identify and shape the elements that have previously complicated optimal implementation of genetics applications in healthcare.

A number of suggestions have been made to explain the relative low implementation rate of genetics, e.g. technology outpacing capacity, lack of organizational structure, absence of a collective sense of urgency, and disagreement on responsibilities^7–9^. In essence, healthcare is notoriously resistant to change; initiating even the smallest changes necessitates attuning of autonomous actors, consideration of effects beyond healthcare, and compelling evidence to break with tradition^10^. On hindsight, envisioning a complete overhaul kickstarted by the mere publication of the human reference genome – common sense in the early 2000s – may have been somewhat optimistic.

Still, various transitions in systems similar to healthcare have (recently) occurred, with the energy transition as a prime example^11–15^. These transitions are characterized by two success factors. First, the focus is on sustainability, not only thematically (e.g. environment, waste, climate), but also in how technology was implemented (e.g. roof-top solar panels and direct-to-consumer (online) selling of electric cars). The transition drivers (unconsciously) adhere to Amara’s Law, which states that “we tend to overestimate the effect of a technology in the short run and underestimate the effect in the long run”^16^. Second, there was a growing understanding of the dynamics at the various system levels, e.g. on how sub-domains or regimes can become interconnected because of shared niches^17^.

Here we put the developments in human genetics in a transition context. We consider the most prominent sub-domain – clinical genetics – as the central regime, surrounded by multiple *adjacent regimes*^12^. Preliminary explorations identified four genetics-affiliated regimes, with varying degrees of overlap and distinction with respect to structure, culture and practice^9^.

## Materials and methods

We applied the “ambassador model of representation” for collecting input from various stakeholders^18^. Briefly, we interviewed representatives of each regime or subgroup thereof in homogeneous duos or trios. From each interview group we invited one representative (‘ambassador’) to participate in the multidisciplinary focus group (**Figure 1**).

**Figure 1.**
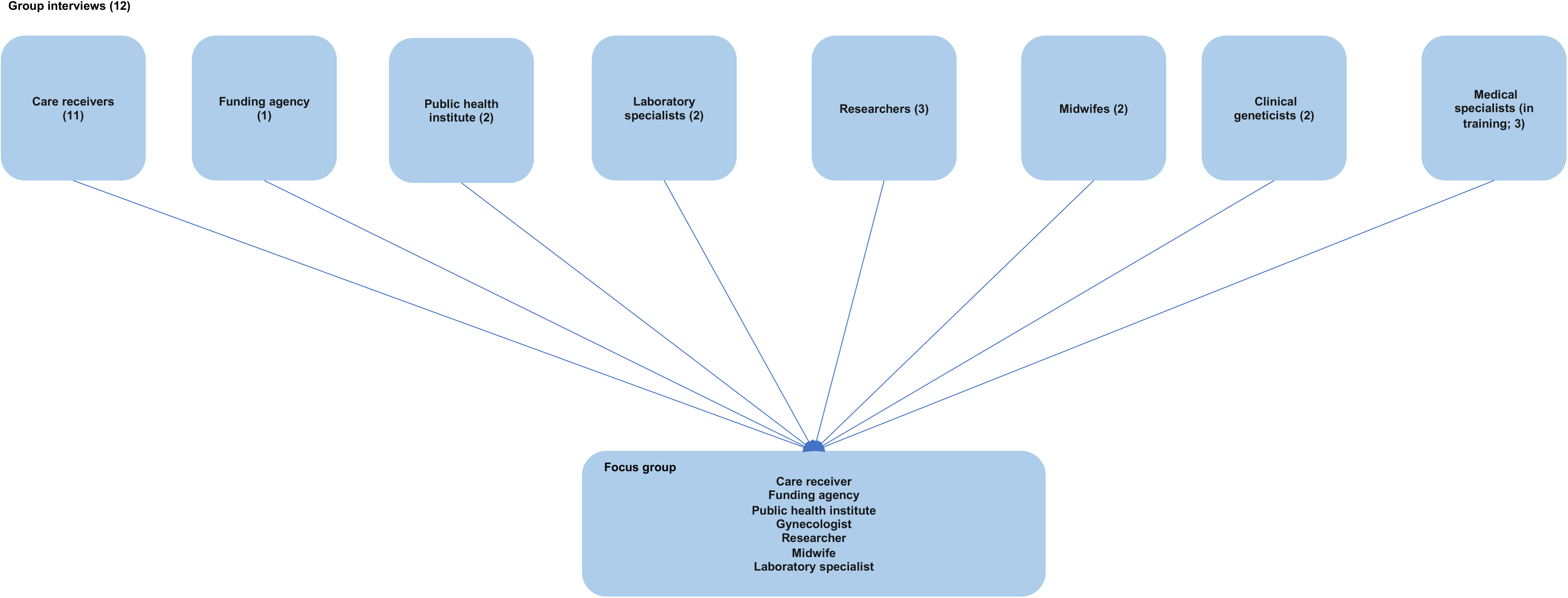
Participants and study design. Nine actor groups were included, and initially interviewed in homogeneous group interviews. From each of the 9 actor groups, one representative (‘ambassador’) was invited to participate in the subsequent focus group.

### Regime descriptions

Based on an initial literature scan, we identified four regimes that were closely connected – partly entangled – with clinical genetics. We collected key characteristics of four adjacent regimes – public health, human genetic research, primary and secondary healthcare, and society - based on scientific literature and publicly available information.

### Participants

Participants were recruited using purposeful sampling on the basis of the actor groups: laboratory specialists (LS1-LS3), clinical genetics (CG1-CG2), researchers in human genetics (HGR1-HGR3), non-genetic medical specialists (ngMS1-ngMS3), midwives (MW1-MW2), care receivers (citizens and patients; CR1-CR11), and representatives from a public health institute (PH1-PH3) and a funding agency (FA1). Participants of the focus group were recruited during the group interviews. All participants consented for data collection and publication.

### Group interviews

To explore the perspectives of the different actor groups, we conducted twelve semi-structured face-to-face homogeneous group interviews (n=2-4 per group, 26 in total: see figure 1 for details). Homogenous participant groups of all relevant actors provided us with profession-specific perceptions of the regime and also enabled elaboration on expectations of other actor groups freely, without interference from other actors. All interviews were conducted in the native language (Dutch). Most interviews lasted approximately 60 minutes.

### Transcription and analysis

All interviews were transcribed verbatim and coded using a coding tree. To help closed coding, the data analysis software ATLAS Ti was used. In addition to this closed coding, a summary was made to ensure interesting quotes that did not belong in the coding tree were captured (open coding). This was also used to assemble quotes for one aspect from various actor groups in one document (axial coding). All coding was conducted by a single researcher to ensure consistency.

### Focus group

Most striking themes from the interviews gave guidance in the design of the focus group. This entailed exercises to elaborate on: 1) what these actors exchanged; 2) the General Data Protection Regulation; 3) technologies; and 4) which patients they helped. The focus group lasted approximately three hours. The focus group was recorded and transcribed verbatim. Selective coding was conducted based on preliminary results from the group interviews. An observer took notes simultaneously to document non-verbal interaction such as nodding to indicate approval or disagreement, facial expressions, and sphere. Two observers interpreted discussions and summarized the most important conclusions.

## Results

Starting from clinical genetics as a distinct regime, we explored four adjacent regimes – public health, human genetics research, non-genetic healthcare, and society (**Figure 2**). Literature and public information provided the key characteristics of each adjacent regime. The group interviews and subsequent focus group provided deeper insight in the structure, culture and practice of each regime, as well as determinants for adoption of genetics applications in adjacent regimes.

**Figure 2.**
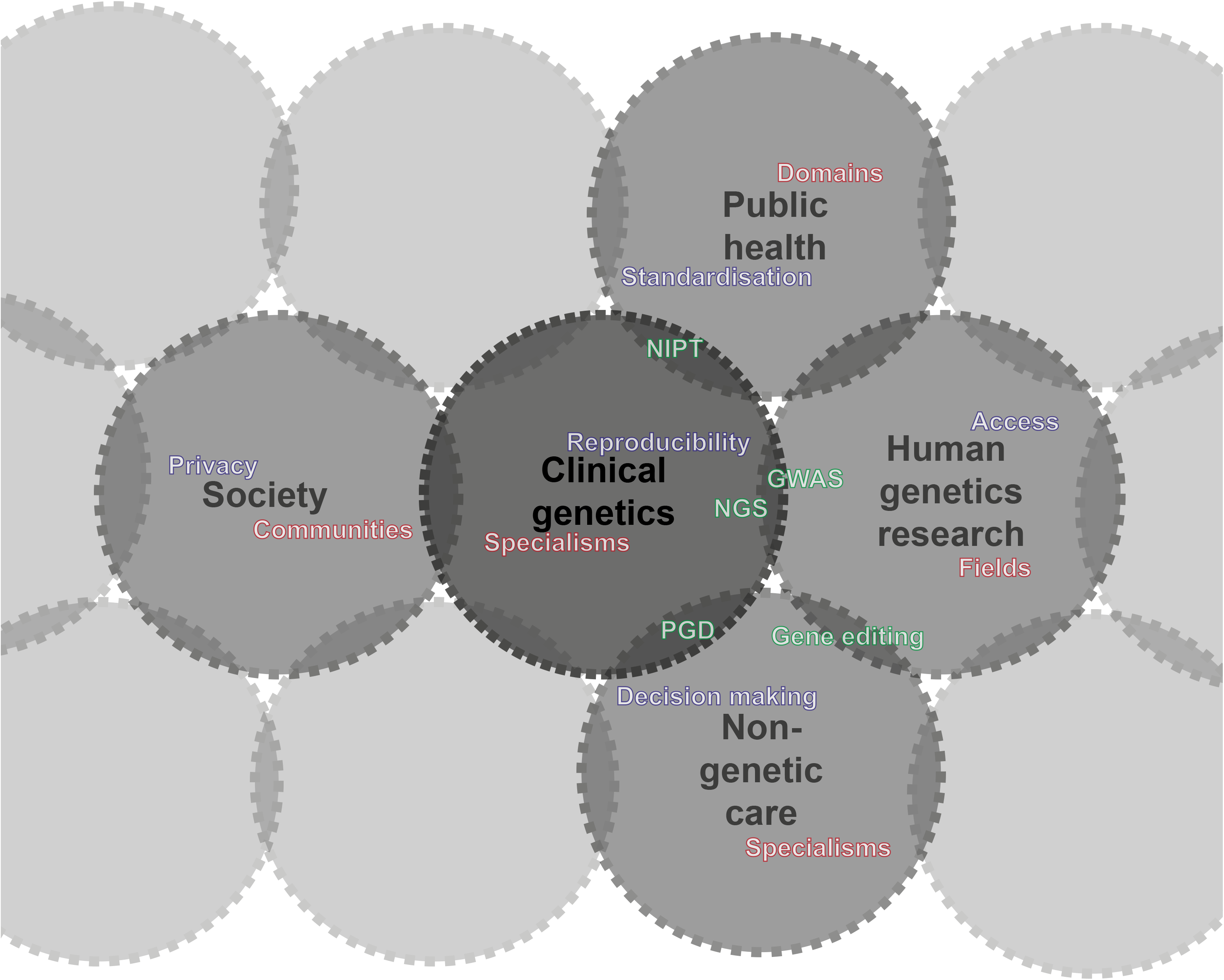
Clinical genetics and adjacent regimes. Starting from clinical genetics as the regime under study (central circle), the adjacent regimes (dark grey) are public health, human genetic research, non-genetic care and society. Other regimes (light grey) affect clinical genetics mildly or indirectly, and are therefore not part of this study. The interaction between clinical genetics and adjacent regimes is determined by the perception on data use (blue), expectations from shared technologies (green) and the discriminator for compartimentalisation (red). NGS = next-generation sequencing; GWAS = genome-wide association studies; NIPT = non-invasive prenatal testing; PGD = pre-implantation genetic diagnostics. Adapted from Van Raak *et al*.^12^

### Public health

Literature shows that public health emerged in the 1920s – simultaneous to modern medicine – predominantly as a result of massive industralisation and urbanization^12^. It is generally defined as "the science and the art of preventing disease, prolonging life and improving quality of life”^19^. Vaccination, environmental safety, and lifestyle interventions are hallmark public health services, which typically require national, coordinated action to ensure fully distributed implementation. Or, as Participant PHI1 put it: “[public health institutes] *are amidst the triangle of practice, policy and science*”. Public health programs in the Netherlands are usually upon directive from the Ministry of Public Health, Welfare and Sports (VWS), and thus embedded in a structure of national regulation and financing, a culture of prevention, and a practice at the local level (**Table 1**).

**Table 1.**
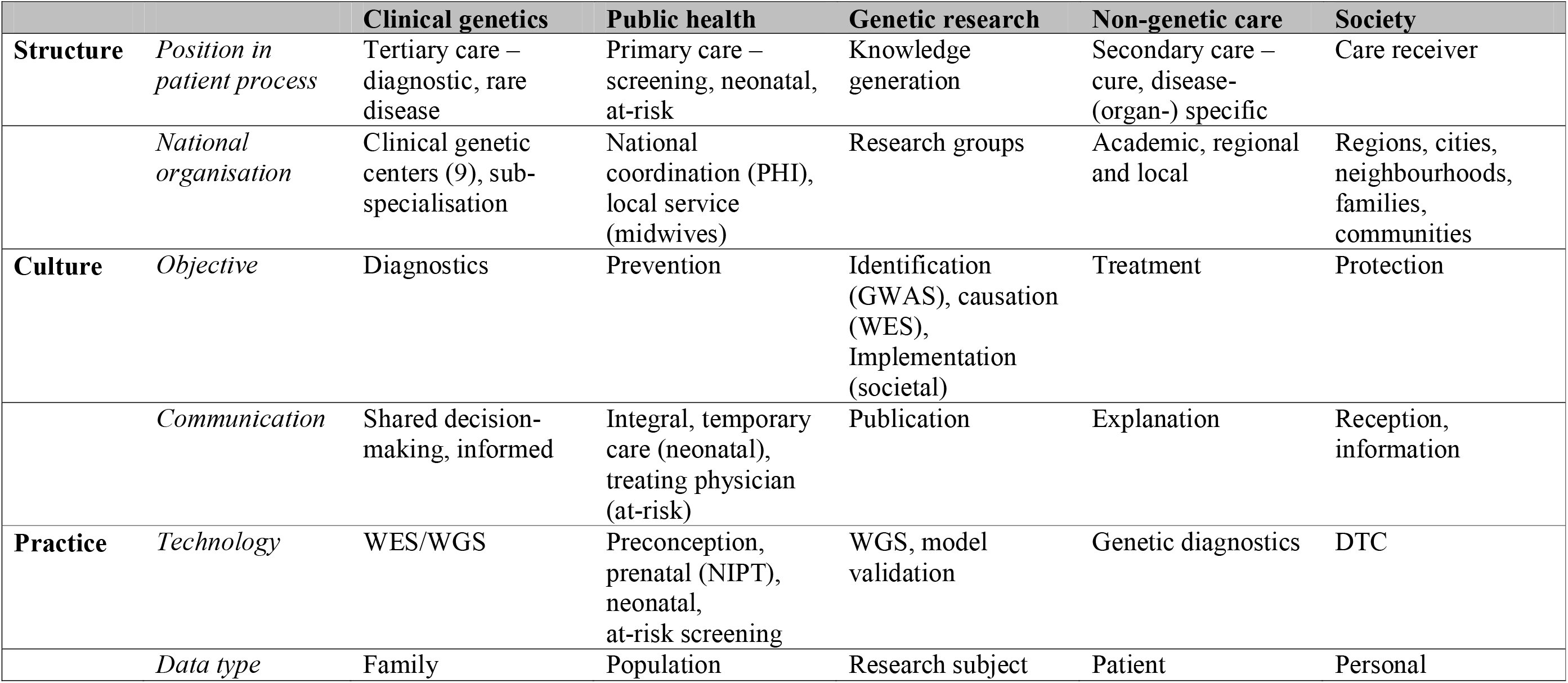
Regime characteristics of clinical genetics and its adjacent regimes.

The most relevant public health service in the context of clinical genetics is (preventive) population screening. This increasingly involves some form of genetic testing, either as part of the first-tier test (e.g. neonatal screening) or as a follow-up (e.g. breast cancer screening)^20^. Whereas coordination of such programs is the responsibility of the national public health institute, the actual testing is generally performed by ‘field parties’. For instance, neonatal screening is typically initiated by midwives and youth healthcare physicians, while pediatricians and clinical geneticists take over if follow-up is required^21^. Similarly, the national screening programs for breast, colorectal and cervical cancer occur under auspices of the national public health institute, but the actual screening occurs at dedicated screening centers. Other forms of genetic screening (e.g. preconception or pharmacogenetic), are not (yet) fully established at the population level and offered on occasional basis^22,23^.

### Human genetics research

Most innovations in clinical genetics have sprouted from human genetics research, a scientific discipline with roots in the nineteenth century (Darwin, Mendel), which has vastly matured since the publication of the human reference genome^3,4,24^. Notwithstanding the emerging wave of applications to *edit* particular genetic loci (i.e. through CRISPR/Cas), the majority of human genetics research has been on the *identification* and *understanding* of disease loci. Following the human reference genome publication, large efforts were undertaken to map human genetic variation, and search for genetic variants underlying disease. In particular the quest for disease loci divided the field; many research groups performed large-scale genome-wide association study (GWAS) on common disease (e.g. cancer or cardiovascular disease), while others performed smaller-scale mutation-detection studies in patients with rare (syndromic) diseases (e.g. intellectual disability or congenital disorders)^25,26^. In total, more than 50.000+ disease loci have been identified through GWASs, but only few have resulted in a change in medical decision making^25,27^. Simultaneously, many rare diseases have been ‘solved’, but due to their rarity they have only recently started to get recognition as a significant public health issue^28–30^. Recently, the sub-communities within human genetics have started to converge, providing first indications of a new perspective on the ‘evidence base’ that is required for future genetic care^31,32^.

### Non-genetic healthcare

Patients are generally referred to clinical genetics by other – non-genetic – medical specialists, like pediatricians, oncologists, gynecologists, and – occasionally – general practitioners. Secondary healthcare is structured according to specialty, either in a certain organ system (urologist, cardiologist), patient type (pediatrician, gynecologist) type of care (anesthesiologist, surgeon), or disease (oncologist). Historically, medical specialists have evolved out of the physician constellation, and have rivaled with the remaining general doctors (general practitioners) in terms of resources, status and recognition^12^. Since the 1930s, general practitioners have been considered the ‘gateway’ to healthcare, deciding on referral of patients to further specialized care. Since the introduction of the registry of medical specialists in 1973 (BIG), general practitioners have been recognized as a distinct medical specialty, operating in relative autonomy. Growing individualization and a more critical attitude in society at large has undermined the authority of medical specialists (e.g. second opinion, media coverage)^12^. Moreover, the recent trend towards constructing multidisciplinary teams around particular patients has diminished the individual specialist’s autonomy^33^.

### Society

Obviously, society as whole is broader than an individual regime, the actors interacting directly with healthcare could collectively be considered the regime of ‘care receivers’. Traditionally, this would comprise patients, but especially in the case of clinical genetics this may also comprise family members, and healthy individuals at-risk (e.g. in the case of breast cancer). In relation to healthcare, there is a great need for responsible use of personal data. In Europe, this has culminated in the European General Data Protection Regulation (<u>GDPR</u>). Meanwhile, the idea of what privacy entails is changing under the influence of internet and communication technologies, which poses an additional layer of complexity to e.g. the implementation of Electronic Health Records (EHR). Long-existing medical regulations (e.g. in the Netherlands Article 2 of the Law on Special Medical Operations – Wbmv – and the Population Screening Act – Wbo) have come under pressure as a result of developments in medical technology and the perception of ‘good care’. Finally, there is a society-wide tendency for back-to-nature and healthy lifestyle products and a decreasing support for (public) funding for (genetic) research, thus requiring novel business models to finance healthcare innovation.

### Cross-over of genetics in adjacent regimes

The insights in the structure, culture and practice of the four regimes adjacent to clinical genetics logically led to reflections on the impact of genetic applications on each regime, and on the determinants for application of emerging genetic technologies. Three aspects were frequently addressed as determinants for application: the use of data, the expectations from technology and the level of transdisciplinary empathy.

> *“Our data generation levels are unsustainable; we simply will not be able to store all data.”*
>
> (Participant LS1)

The actors within clinical genetics (clinical geneticists, laboratory specialists) were mostly concerned with the mere volume of genetic data. Genetic laboratories follow self-imposed national guidelines to mitigate capacity issues (e.g. delete raw data and intermediate analysis files), but still have an annual spending of €50k-€100k on data storage. The *a priori* concern is reproducibility; laboratories need to be capable of reproducing the original diagnosis. Conversely, the actors in adjacent regimes put more emphasis on other dimensions of genetic data. More specifically, each regime seemed to consider genetic data challenging, but for a different reason. For instance, the researchers (human genetics research) experienced challenges with respect to data access and use, particularly since the installment of the Europe-wide General Data Protection Regulation (GDPR). Similarly, clinical geneticists experience the practical implementation of the GDPR as burdensome. It especially causes trouble concerning hereditary data sharing between family members as a consult should be planned for every specific data sharing case. Participants ngMS1 and PHI1 indicated to switch between their roles as respectively healthcare provider and policy maker, and that of a researcher to handle data issue appropriately. Conversely, the care receivers (society) embraced the new data regulations as welcome safeguard for their personal data – not a burden. In this context, participant FA1 stressed the importance of standardization, and the efforts required to make data findable, accessible, interoperable and re-usable (FAIR).

> *“In 5 years? You will be considered a fool if you do not have yourself genetically screened prior to reproduction.”*
>
> *(Participant ngMS1)*

Clinical genetics is largely technology-driven, fed by developments in human genetics research. Whereas the majority of genetic technologies have been developed to *detect* genetic variation, much of the recent attention has gone to CRISPR/Cas, a technology that provides a way to *alter* genetic variants. This affects the perception of genetic applications in society, but also in the other adjacent regimes. For instance, the gynecologists and midwives (primary and secondary care) anticipate a demand for ‘designer babies’ in their practices, which – in their view – is the logical next step after non-invasive prenatal testing (NIPT), pre-implantation genetic diagnostics (PGD) and screening (PGS) and preconception carrier screening. Similarly, the program coordinator (funding agency) indicated to have set up programs and work groups to explore applications of somatic gene therapy. Remarkably, the perception of what was considered ‘technology’ varied considerably between clinical genetics and adjacent regimes. A novel technology in clinical genetics is generally linked to laboratory instruments (e.g. whole-exome sequencing (WES), whole-genome sequencing (WGS) or CRISPR/Cas) which can be used for various applications. Conversely, actors in primary and secondary care, public health, and society referred to the applications as ‘novel technologies’. For instance, non-invasive prenatal testing (NIPT) is considered a technology in gynaecology and obstrectics, while the actual testing comprises a particular form of next-generation sequencing (NGS), which is considered the technology in clinical genetics and human genetics research. Similarly, NGS is the laboratory technology underlying ‘gene panels’ and ‘screening’; the emerging technologies in the eyes of the medical specialists in training (**Figure 2**).

Simultaneously, technology may be a determinant for the level of control and autonomy. Participant PHI1 indicated that the decision to invest in building in-house technological infrastructure strongly depends on the need to build independent expertise. Similarly clinical geneticists and laboratory specialists are strict on quality, reliability and reproducibility, and having in-house technology increases control over these aspects. Conversely, the focal point of other medical specialists is on control over treatment decisions, and thus on being the treating physician. This illustrates mixing genetic implications is perceived tricky, especially when another specialized doctor is the treating physician of a patient with clinically relevant genetic aspect.

> “*Every field has its own methods*.”
>
> (Participant HGR1)

Medical specialists call it ‘specialties’, researchers talk about ‘fields’, and policy makers refer to ‘domains’. They all refer to the same phenomenon; their regime is structured into compartments that operate in relative independence and isolation. The compartments in clinical genetics are subspecialisations, e.g. oncogenetics, cardiogenetics, or neurogenetics. Adjacent regimes are also highly siloed, but the determinants of separation vary considerably between regimes. The subspecialisation in clinical genetics originates from the silos in general healthcare, where doctors specialize in particular organ systems (e.g. urologist, cardiologist), patient types (e.g. pediatrician, geriatrician), type of care (e.g. gynaecologists, anesthesiologists), or disease (e.g. oncologist, syndromologist). The emergence of multi-disciplinary teams increase permeability of silo walls, but it forces the specialists to gather at the same (virtual) location at the same time, which can be challenging. Still, the clinical geneticists consider the multidisciplinary teams as a sustainable future to withhold patients from ‘shopping around’ with their own stories.

Human genetics research knows numerous fields, often linked to distinct disease categories or biological concepts. The scientists considered commitment to a particular field as an important factor in their career development; it provides visibility and a clear profile, which is beneficial in the grant review process, where reviewers may think: ‘Yeah, that and that researcher, working on that and that’. On the other hand, if researchers’ focus is too narrow, it would slow down their research. Yet the main discriminator of silos is inherent to the culture of science, and concerns the objective and corresponding research approach and methodologies, with two major silos as a result; fundamental research and translational research. The scientists referred to the aim as being the essence of the distinction between fundamental and translational research, and its importance in finding sustainable healthcare solutions. Fundamental research – as they see it – is meant to *understand* diseases, symptoms and biological processes. The next step would be to *translate*, e.g. by predicting response of an individual patient to a particular treatment. While hard criteria for what makes a researcher fundamental or translational do not exist, the key seems to lay in the primary motivation. Of the three scientists we interviewed, one indicated to be primarily curiosity-driven, one preferred to do research with direct benefit for patients, and the third valued both motivations equally.

Simultaneously, the increasing societal demand for instant revenues from public investments in scientific research was recognized, especially among policy-oriented actors, i.e. from funding agencies and public health institutes. The stakeholders generally navigate between silos in policy, science and society, which can be challenging. For instance, participant PHI1 mentioned neonatal screening as an example where the policy area formally is prevention, but relevant counseling occurs in a context of medical care.

## Discussion

Based on twelve group interviews and a focus group with genetics-affiliated professionals we explored structure, culture and practice in four adjacent regimes of clinical genetics. Three elements require specific attention when implementing genetic applications in adjacent regimes – data, technology and silos.

Actors often point to structure – predominantly regulation – as major obstacle for implementation of novel genetic application, while the underlying issue is often one of culture. This is clearly illustrated – in this study and in literature – by the example of General Data Protection Regulation (GDPR). Genetics professionals often perceive the GDPR as a burden on their clinical or research practice, implying that the discrepancy between the regime’s structure and practice was caused by the regulatory newbie^34^. Yet the evolving societal attitude towards data use – as expressed by the citizens in our study – indicates a necessity to reconsider the existing culture (and resulting practice) towards use of genetic data.

Similarly, the discriminators of silo-formation (structure) are largely defined by culture. Clinical genetics is subspecialised according to the disease fields and organ systems that have prevailed in modern medicine. The introduction of clinical genetics in the 1970s – when the healthcare system had been largely institutionalized – forced the new medical specialty to fit in the structure of the healthcare regime^12^. This has resulted in a culture of sub- or even hyperspecialisation in clinical genetics. Conversely, the non-medical adjacent regimes are also siloed, but in different directions. Public health is divided in curative and preventive care, and human genetic research is predominantly divided into fundamental and translational research.

Finally, the cultural discrepancies were even apparent from the perception of technologies. Where researcher and lab analysists name underlying technologies of genome care, care providers and -receivers address the care application.. Researchers were not aware of the clinical implication of research; care providers were not aware of underlying technologies of the clinical application nor of upcoming technologies.

Although generally not considered a regime in itself (landscape), the organisation of funding and the role that funding agencies have in the distribution of resources strongly affects the implementation of clinical genetics in adjacent regimes. For instance, research grants are largely awarded based on publication records of individual researchers, which is difficult to align with the activity-based reimbursement schemes in clinical genetics and other healthcare regimes. Similarly, while alternative financing schemes are available for exceptional services in healthcare (e.g. expensive medicine, in-vitro diagnostics), no such alternatives exist for clinical genetics, hampering e.g. the application of whole-genome sequencing for multiple diagnostics or prognostic purposes.

This study provides nuances to the observation that clinical genetics lacks organizational structure, absence of agency, and attuning of stakeholders^8,9^. We acknowledge that an overarching organizational structure and alignment is lacking, but this is rather due to the lack of alignment between clinical genetics and its adjacent regimes, than to an absence of structure. Therefore, attuning of stakeholders and creating a shared vision is important^9^. To reach this shared vision, it is essential to first acknowledge that each investigated regime not only comprises genetic applications, but entails their own, non-genomic focused, characteristic regime in abundance. We therefore recommend to be sensitive for the differences with adjacent regimes, instead of trying to reshape these regimes, e.g. through change agents^9^.

To conclude, the genome care regime cuts through the adjacent regimes of care, research, institutions and care receivers. Each regime comprises signature cultural, technological, industrial aspects. Transdisciplinary empathy towards adjacent regimes on those aspects could lead to a more comprehensive regime of genome care.

## Data Availability

Transcripts are available with the corresponding author upon request

## Notes

### Competing Interest Statement

The authors have declared no competing interest.

### Funding Statement

No external funding was received

### Author Declarations

METC Utrecht confirms that the Medical Research Involving Human Subjects Act (WMO) does not apply to this study (reference: WAG/mb/20/020940) METC Utrecht Heidelberglaan 100 3584 CX Utrecht T: +31887556376 E:

